# Exploring the use of preprints in dentistry

**DOI:** 10.1101/2023.07.11.23292516

**Authors:** Rafael Sarkis-Onofre, Carolina Girotto, Bernardo Antonio Agostini

## Abstract

**Objective:** This study aims to assess the use, impact, and dissemination of preprints in dentistry.

**Methods:** This is a meta-research study with a cross-sectional design. We included preprints published in dentistry, regardless of the year of publication. Searches were performed in the medRxiv.org and Preprints.org platforms and restricted to English. One researcher extracted the data, and another researcher verified data consistency. The following data were extracted: year of publication, country of the corresponding author, number of abstract and full-text views and downloads, Altmetric attention score, whether the preprint was mentioned in other servers such as Twitter and Publons, number of mentions in other servers, number of citations in the Dimensions database, and whether the preprint had already been published in a peer-reviewed journal. If already published, we extracted the journal’s impact factor (JCR 2021) and the number of citations in the Dimensions database. We conducted a descriptive analysis of the extracted characteristics and explored relationships between metrics using the Spearman correlation.

**Results:** We identified 276 preprints. Most of the studies were published between 2020 and 2022 (*n* = 229), especially those from ten countries. The most-cited preprint and published article are the same study. Only the correlation between the number of preprint citations and peer-reviewed article citations in the Dimensions database showed a large positive association (Spearman’s rho = 0.5809).

**Conclusion:** Preprints gained popularity over the last several years due to the COVID-19 pandemic and reached a larger audience, especially on platforms such as Twitter.

**Clinical Significance:** Preprint publishing allows faster dissemination of science for the benefit of society.

## Introduction

Traditionally, research has been communicated through scientific journal articles. The publishing process involves many steps, including peer-review of the manuscript by other researchers [1]. Bjork et al. [2] highlighted that the time between submission and article acceptance can be as long as six months; however, average publication times in disciplines such as business and economics can be longer. This long publication process might not provide the prompt, science-based solutions needed for real-life problems. Additionally, in critical global emergencies, such as the COVID-19 pandemic, faster responses by the academic community are necessary, and, in that scenario, preprints could be essential[3].

Preprints are initial versions of research manuscripts that have not been peer-reviewed and are publicly available through various platforms, such as medRxiv.org, Preprint.org, and bioRxiv.org [3]. Preprints are not a new publishing format and have been used for over 30 years, but their use has increased recently [3], mostly due to COVID-19. Fraser et al. determined that more than 30,000 research articles related to COVID-19 were posted on preprint servers [3]. The main benefit of preprint publishing is the possibility of accelerating the use of science for the benefit of society[4]. However, widespread use of preprints could involve sharing articles with methodological flaws or biases[5].

As the use of preprints has increased, many discussions have occurred in the literature about the advantages and disadvantages of preprints [4–7]. In dentistry, no study to date has explored the use of preprints. Thus, our objective is to assess the use, impact, and dissemination of preprints in dentistry.

## Material and methods

### Study design and protocol

This is a meta-research study with a cross-sectional design. The study protocol is available through the link https://osf.io/6hpdj.

### Eligibility criteria

We included preprints published in dentistry and oral medicine, regardless of the year of publication. We used the definitions of dentistry and oral medicine as determined by the preprint platforms[8,9]. Preprints were considered preliminary reports of work that have not undergone peer review.

### Search and screening

Searches were performed in the medRxiv platform, a preprint server for health sciences, through the subject area “Dentistry and Oral Medicine” and the Preprints platform through the subject area “Dentistry.” The medRxiv platform allows authors to post research articles, systematic reviews and meta-analyses, clinical research design protocols, and data articles. The Preprints server publishes articles in the following categories: article, review, conference paper, data descriptor, essay, brief report, case report, communication, short note, technical note, and hypothesis. We did not restrict our search based on date; however, only articles published in English were considered. Searches were performed in November 2022.

After identifying preprints in these platforms, all records were uploaded to Mendeley reference management software.

### Data extraction

First, we selected ten studies for a pilot test of data extraction to ensure consistency in interpretation of items. During the pilot test, the reviewers involved in this phase of the study discussed all data to be extracted. Subsequently, one reviewer extracted the data, and another reviewer verified data consistency. In the case of a doubt or inconsistency, the data were extracted again. The following data were extracted: year of preprint publication, country of the corresponding author, number of full-text views and downloads, Altmetric attention score, whether the preprint was mentioned in other servers such as Twitter and Publons, number of mentions in such servers and in which platform the preprint was mentioned, whether the preprint had already been published in a peer-reviewed journal (based on the preprint server information), and number of citations in the Dimensions database. If already published, we also extracted the number of citations of the article in the Dimensions database and the journal’s impact factor (JCR 2021). We chose the Dimensions platform because it includes preprints from several sources (including medRxiv and Preprints) and their respective citation counts. Data extraction was performed in November 2022.

Finally, during data analysis, we decided to collect the Altmetric score and the number of mentions on other platforms for the ten most-cited articles to determine whether the attention received in an academic setting (through the number of citations) was comparable to that received on online platforms, such as Twitter and Publons.

### Data analysis

We conducted a descriptive analysis of the extracted characteristics. Categorical variables are presented as proportions, and numerical variables are summarized by their means and standard deviations. In addition, we assessed the most-cited preprints and articles in the Dimensions database as well as their Altmetric score and the number of mentions in other servers such as Twitter and Publons. The Shapiro-Wilk test was used to assess the normality of numeric data. We performed Spearman correlation tests to determine the relationship between the number of peer-reviewed article citations in the Dimensions database and the following: the number of preprint citations in the Dimensions database, the number of preprint views (the number of full-text views and downloads), the preprint Altmetric score, and the number of mentions on other servers. The analyses were conducted using Stata 14.0 software (Stata Corp., College Station, TX, USA).

## Results

Table 1 summarizes the characteristics of the preprints reviewed. We identified 276 preprints: 131 published in the Preprints server and 145 in the medRxiv server. The list of included preprints is presented in the Supplemental Material. Most of the preprints were deposited between 2020 and 2022 (*n* = 229). Ten countries produced 60.14% of preprints; the USA produced the largest number, with 43 preprints (15.58%), followed by the United Kingdom, with 25 preprints (9.06%). Forty countries published three or fewer preprints, for a total of 60 (21.74%). One hundred thirty-five preprints (48.9%) were subsequently published in peer-reviewed journals, most often in 2021 and 2022 (*n* = 92; 65.15%).

**Table 1:** Characteristics of included preprints.

| **Server** | **n** | **%** |
| --- | --- | --- |
| Preprints | 131 | 47.46 |
| medRxiv | 145 | 52.43 |
| **Year of preprint publication** |  |  |
| 2016 | 2 | 0.72 |
| 2017 | 4 | 1.45 |
| 2018 | 22 | 7.97 |
| 2019 | 19 | 6.88 |
| 2020 | 85 | 30.8 |
| 2021 | 66 | 23.91 |
| 2022 | 78 | 28.26 |
| **Country** |  |  |
| 40 countries with 3 or less preprints | 60 | 21.74 |
| USA | 43 | 15.58 |
| UK | 25 | 9.06 |
| Italy | 20 | 7.25 |
| India | 14 | 5.07 |
| Brazil | 13 | 4.71 |
| China | 12 | 4.35 |
| Japan | 10 | 3.62 |
| Portugal | 10 | 3.62 |
| Spain | 10 | 3.62 |
| Iran | 9 | 3.26 |
| Germany | 8 | 2.90 |
| Chile | 7 | 2.54 |
| France | 6 | 2.17 |
| Saudi Arabia | 6 | 2.17 |
| Lebanon | 5 | 1.81 |
| Slovakia | 5 | 1.81 |
| Taiwan | 5 | 1.81 |
| Egypt | 4 | 1.45 |
| United Arab Emirates | 4 | 1.45 |
| **Preprints published in peer review journals** | 135 | 48.9 |
| **Year of article publication** |  |  |
| 2016 | 1 | 0.74 |
| 2017 | 4 | 2.96 |
| 2018 | 6 | 4.44 |
| 2019 | 11 | 8.15 |
| 2020 | 21 | 15.56 |
| 2021 | 48 | 35.56 |
| 2022 | 44 | 32.59 |

Table 2 presents the metrics related to preprints and peer-reviewed journal articles. Preprints received a mean of 874 full-text views (SD = 970.7). Most preprints were mentioned on other platforms (*n* = 230; 83.33%), including 214 preprints (93%) that were mentioned at least on Twitter. In addition, the mean number of preprint citations in the Dimensions database was lower than that of article citations in the same database.

**Table 2:** Metrics related to preprints and articles.

|  |  |  |
| --- | --- | --- |
| **Mean of preprint full text views (SD)** | 874 (970.7) |  |
| **Mean of Altmetric score of the preprint (SD)** | 4.1 (22.06) |  |
| **Mean of preprint citations in Dimension database (SD)** | 0.75 (1.96) |  |
| **Mean of article citations in Dimension database (SD)** | 9.45 (20.16) |  |
| **Mean of number of mentions of the preprint in other platforms (SD)** | 8.2 (36.6) |  |
| **Mentions** | **n** | **%** |
| **Preprint mentioned in other platform** | 230 | 83.33 |
| **Platforms\*** |  |  |
| Twitter | 214 | 93 |
| Mendeley | 66 | 28.7 |
| Peer review sites | 34 | 14.8 |
| Others | 14 | 6.1 |

\*Articles could be mentioned in more than one platform. The percentages are in relation to the number of the preprints mentioned in other platforms.

Table 3 lists the most-cited preprints and peer-reviewed articles. The most-cited preprint and most-cited article are the same study, which assessed saliva as a candidate for COVID-19 diagnostic testing [10]. In addition, five preprints included in this list of most-cited preprints are also included in the list of most-cited peer-reviewed articles [10–14]. Most of the articles included in both lists are COVID-19–related studies. The most-cited preprints and articles did not necessarily have higher Altmetric scores or mentions on other platforms.

**Table 3:**
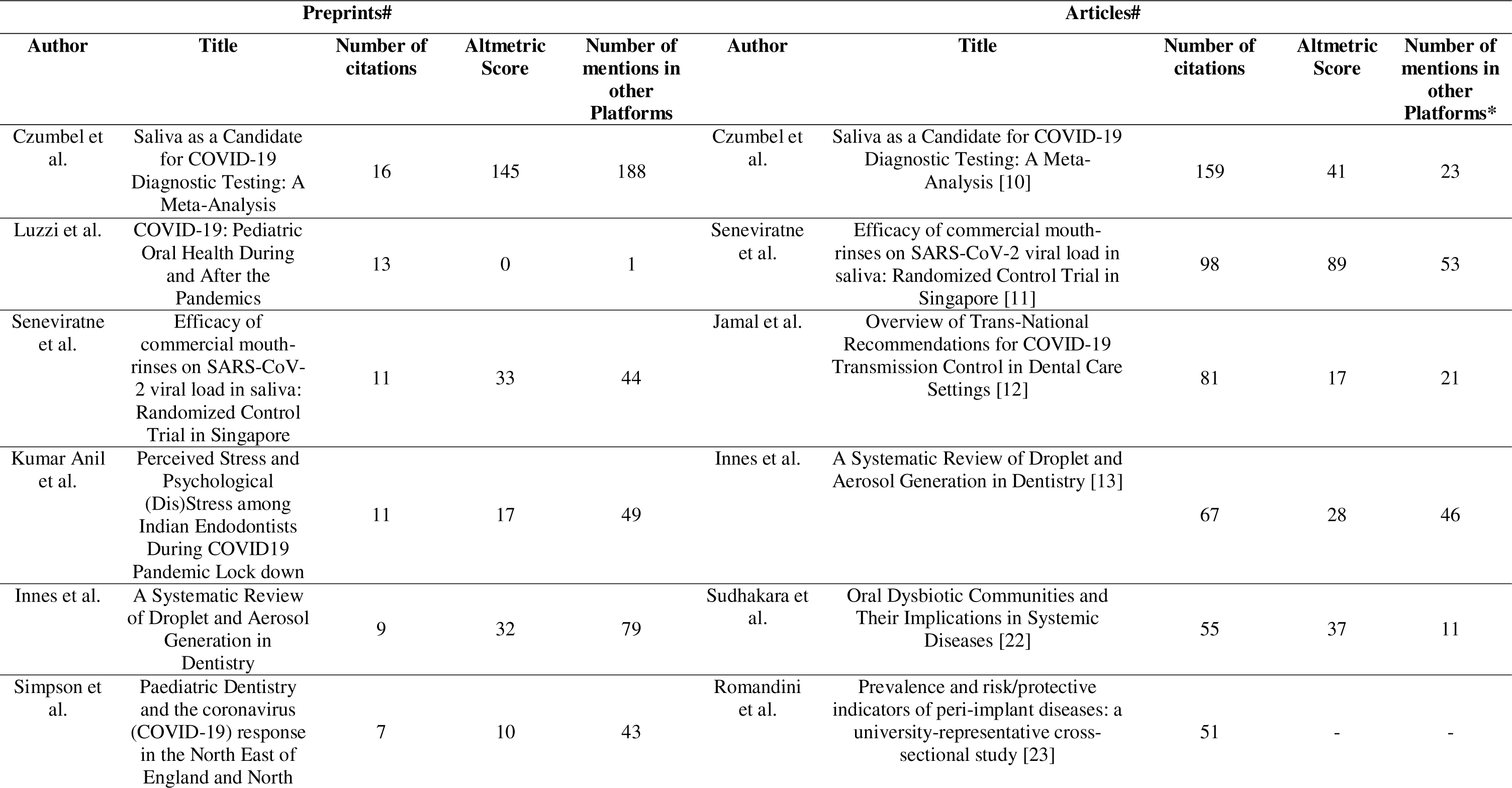
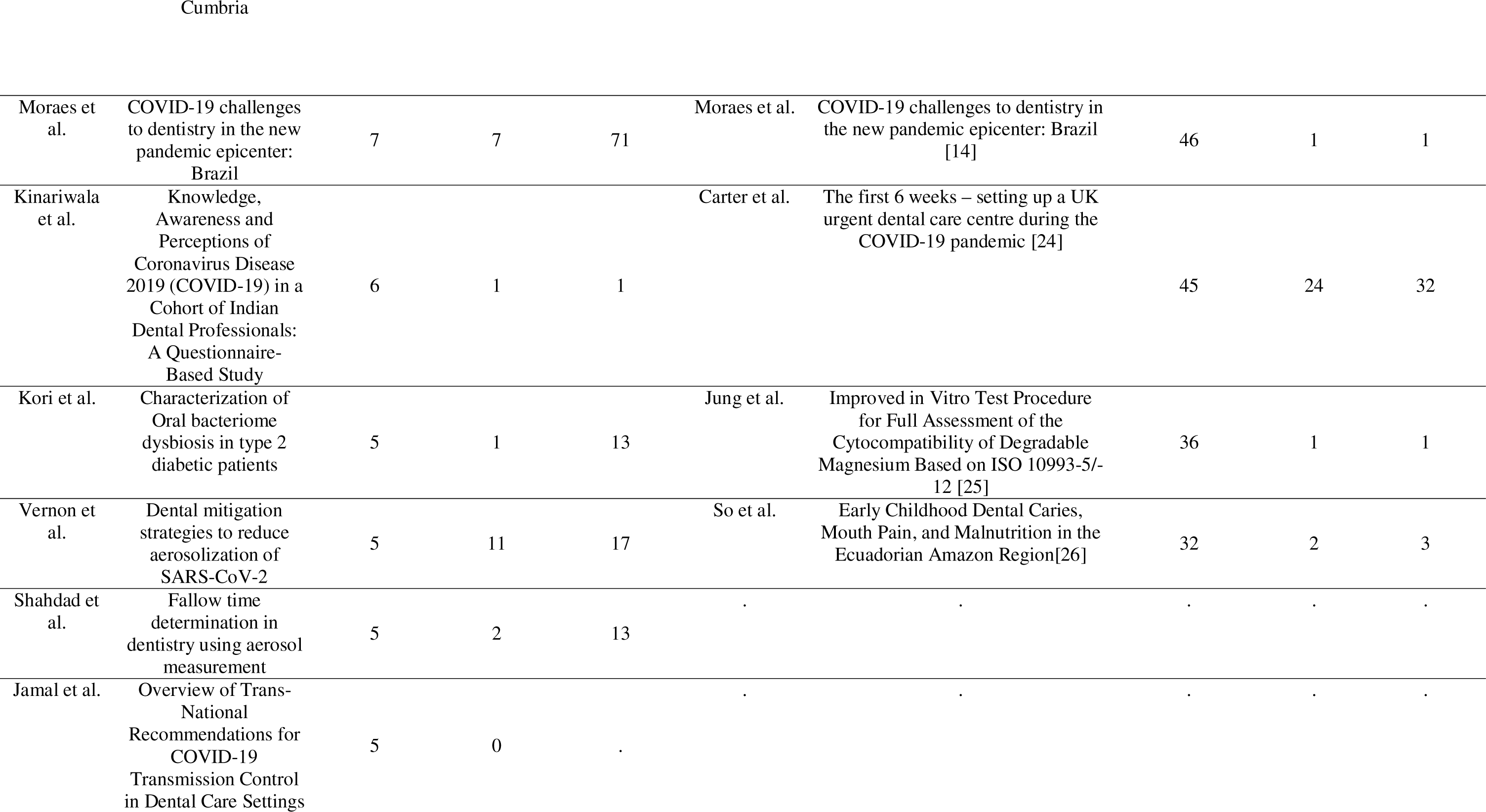
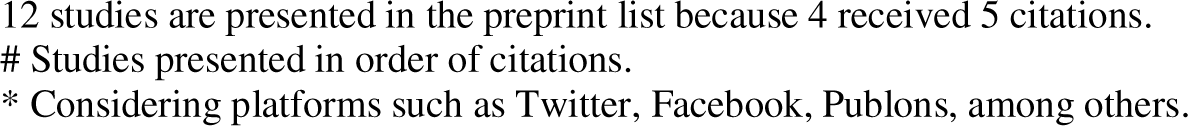
Top cited preprints and articles and related Altmetric scores and number of mentions in other platforms Preprints# Articles#.

Table 4 presents the results of the correlation tests. Only the relationship between the number of preprint citations in the Dimensions database and the number of peer-reviewed article citations in the same database showed a large positive association (Spearman’s rho = 0.5809).

**Table 4.**
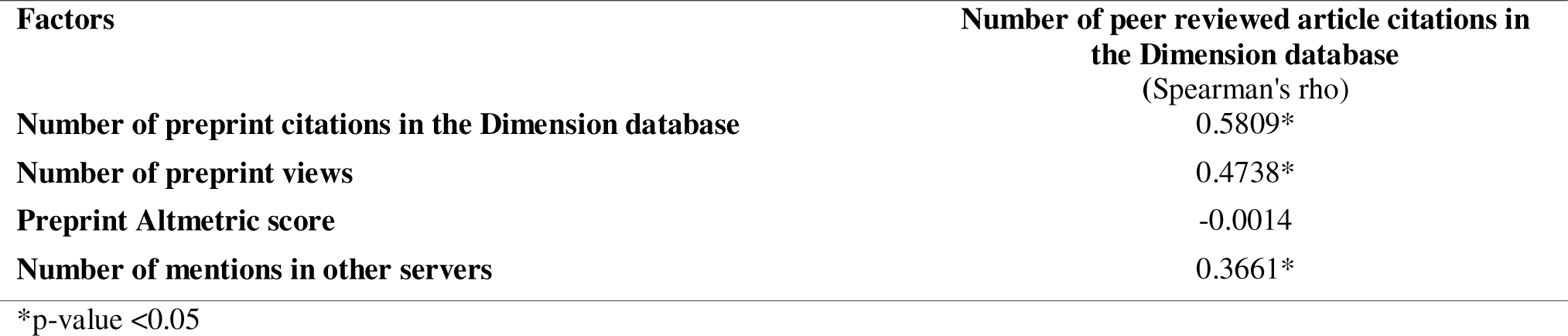
Results of correlations tests between preprint and article metrics.

| **Factors** | **Number of peer reviewed article citations in the Dimension database**<br>**(Spearman's rho)** |
| --- | --- |
| **Number of preprint citations in the Dimension database** | 0.5809* |
| **Number of preprint views** | 0.4738* |
| **Preprint Altmetric score** | -0.0014 |
| **Number of mentions in other servers** | 0.3661* |

\*p-value <0.05

## Discussion

This is the first study to explore the use of preprints in dentistry. The importance of preprint publishing has been increasing over the last several years due to the need to accelerate the application of science to benefit society, especially during the COVID-19 pandemic. Considering the widespread use of preprints, it is important to understand the impact of this method of publishing on a dentistry science audience. Some findings are important to highlight: (1) preprints gained an audience on platforms such as Twitter, 1. (2) studies subsequently published in peer-reviewed journals tended to receive more citations than they did in preprint format, (3) the most-cited preprints and articles did not necessarily gain more attention on other platforms, and (4) the number of preprint citations could be an indicator of future article citations.

Rapid dissemination of information seems to be the main benefit of preprints [15]. In this context, sharing preprints on Twitter could engage a larger audience [3]. Indeed, we found that most preprints we reviewed were mentioned on Twitter, and the mean number of mentions on other platforms (such as Twitter and Publons) and the Altmetric score were 8.2 and 4.1, respectively. In addition, the most-cited preprints and articles did not necessarily earn higher Altmetric scores or mentions on other platforms, demonstrating that the interests of scientists and the broader public might differ. Fraser et al. found similar results when considering preprints of COVID-19 research in the first ten months of the pandemic [3]. Furthermore, three recent studies have revealed that journal articles that were previously posted on preprint servers received more attention, in terms of citations and Altmetric scores, than articles not previously posted, highlighting another advantage of the use of preprints [16–18]. Finally, it is important to emphasize that a high number of citations does not necessarily indicate high quality [19].

The lack of quality assurance is the main concern associated with preprints, especially because no peer-review process is involved [15]. As would be expected given this concern, we found that, while the numbers of preprint citations and peer-reviewed article citations were significantly positively correlated, the mean number of preprint citations was lower than that of article citations. In other words, scientists prefer to cite published articles over preprints, maybe due to the benefits of peer review. However, we should note that there is a slight quality increase between preprints and the subsequent peer-reviewed articles [20], and the current peer review process does not ensure the publication of high-quality articles [19]. Furthermore, there is an ongoing discussion of whether the peer-review process actually produces higher-quality science [19].

Another critical barrier to using preprints is a lack of knowledge about the process [21]. In a recent study, a high number of survey respondents indicated that they did not post research results in preprint servers because they did not know it was a possibility[21]. The study found that this response was most common among female authors, early-career researchers, and non-US-based researchers[21]. These findings highlight the importance of our study and the need for better communication about the advantages of preprints.

We did not explore preprints separately by subject; however, we noted that preprints gained popularity over the last several years. This increase is likely due to the COVID-19 pandemic, as evidenced by the prevalence of studies about COVID-19 among the most-cited preprints and articles and by the publication of most of the preprints between 2020 and 2022, during the pandemic. Fraser et al. highlighted the importance of preprints in disseminating knowledge to academic and non-academic audiences during the COVID-19 pandemic[3].

Our study had several limitations. First, we searched only two preprint databases and included only articles published in English, so the results may not be generalized to other databases and languages. Second, we did not perform the data extraction in duplicate; however, a pilot test was performed to minimize errors, and one author reviewed possible data inconsistencies. Third, we did not assess the preprints based on subject because our goal was to gain a broad understanding of preprints in dentistry.

Finally, more studies assessing the barriers preventing researchers from posting preprints and the attitudes of researchers toward data from preprints are necessary. In addition, it is crucial to consider different research areas and how they might vary in researchers’ attitudes toward and knowledge of using preprints.

## Data Availability

All data produced in the present work are contained in the manuscript

## Acknowledgements

This study is funded in part by the Brazilian Federal Agency for Coordination of Improvement of Higher Education Personnel (CAPES) – Finance code 001. R.S.O is funded in part by Meridional Foundation (Passo Fundo, Brazil). C.G is funded by National Council for Scientific and Technological Development (CNPq – Brazil). The funders had no role in the study design, data collection and analysis, or manuscript publication.

## Conclusion

Preprints gained popularity over the last several years due to the COVID-19 pandemic and reached a larger audience, especially on platforms such as Twitter. In addition, studies published in peer-reviewed journals tended to receive more citations than in preprint format, but the number of preprint citations might predict future article citations.

## List of included preprints

Abe A, Nakayama A, Otsuka Y, Shibata K, Matsui YIto Y, Hayashi H, Momokita M, Taniguchi S. Relationship of preoperative oral hypofunction with prognostic nutritional index in gastric cancer. medRxiv, 2022.

Adegboye A, Santana D, Cocate P, Benaim C, Santos P, Heitmann, Carvalho M, Schlussel M, Castro M, Kac G. Acceptability, Adherence and Retention of a Feasibility Randomised Trial on Periodontal Treatment and Vitamin D/ Calcium Milk Fortification among Pregnant Women: A Mixed-Methods Evaluation. Preprints, 2020.

Adwibowo A. Fuzzy logic assisted COVID 19 safety assessment of dental care. medRxiv, 2020.

Al-Ibrahim I, Alotaibi A, Alshammari A, Madfa A. Experience of Saudi Dental Practitioners with Intraoral Scanners. medRxiv, 2022.

Al-Moraissi E, Abood M, Alasseri N, Günther F, Neff A. Is Standard Personal Protective Equipment Effective Enough To Prevent COVID-19 Transmission During Aerosol Generating Dental, Oral and Maxillofacial Procedures? A Systematic Review. medRxiv, 2020.

Alam M, Ganji K, Alfawzan A, Manay S, Srivastava K, Chaudhari P, Hosni H, Alswairki H, Alansari R. Ectopic Eye Tooth Management: Photobiomodulation / low-level Laser Emission Role in Root Resorption After Fixed Orthodontic Treatment. Preprints, 2022.

Aldayri B, Nourallah A, Badr F. Clinical and Radiographic Evaluation of the Healing after MTA Application on Mechanical Furcal Perforations in Primary Molars-part2. Preprints, 2019.

AlHayyan W, AlShammari K, AlAjmi F, Pani S. The Impact of COVID-19 on Dental Treatment in Kuwait – a Retrospective Analysis from the Nation’s Largest Hospital. Preprints, 2022.

Ali G, Ajili F, Hammami C, Blouza I. Interdisciplinary Management of Radicular Cyst in Anterior Maxilla - A Case Report. Preprints, 2022.

Ali S, Alshabaan S. What Do Parents Know About Oral Health and Care for Preschool Children in the Central Region of Saudi Arabia? Preprints, 2020.

Alkildani S, Mandlule A, Radenković M, Najman S, Stojanovic S, Jung O, Ren Y, Cai B, Görke O, Schmidt F, Barbeck M. In Vivo Analysis of the Immune Response to Strontium- and Copper-doped Bioglasses. Preprints, 2022.

Allison J, Dowson C, Pickering K, Červinskytė G, Durham J, Jakubovics N, Holliday R. Local Exhaust Ventilation to Control Dental Aerosols and Droplets. medRxiv, 2021.

AlMesmar H, AlMashhadani S, Saleh N, Farghali K. The Detection of Pre-Diabetic Patients in the Dental Office. Preprints, 2019.

Almoraissi E, Galvão E, Nikolaos Christidis S, Falci G. Top 100 cited systematic reviews and meta-analyses in the major journals of oral and maxillofacial surgery: a bibliometric analysis. medRxiv, 2020.

Alqalshy E, Ibrahim A, Abdel-Hafiz A, AbdEl-Rahman K, Alazzazi M, Omar M, Abdel-Wahab A. Effect of Docosahexaenoic Acid as a Chemopreventive Agent on Experimentally Induced Hamster Buccal Pouch Carcinogenesis. Preprints, 2022.

Alqerban A. Maxillary Canine Impaction and Unilateral Cleft Lip and Palate: A Review of the Current Literature. Preprints, 2018.

Alves L, Mesaros A, Ponces M, Pollmann M. Aesthetic evaluation of the need for orthodontic treatment – Perception among university students. medRxiv, 2021.

Amitay M, Barnett-Itzhaki Z, Sudri S, Drori C, Wase T, Abu-El-Naaj I, Rieck M, Avni Y, Pogozelich G, Weiss E. Deep convolution neural network for screening carotid calcification in dental panoramic radiographs. medRxiv, 2022.

Ancuceanu R, Anghel A, Ionescu C, Hovanec M, Cojocaru-Toma M, Dinu M. Clinical Trials with Herbal Products for the Prevention of Dental Caries and Their Quality: A Scoping Study. Preprints, 2019.

Aoun G, Aoun P, El-Outa A. Anemia: Considerations for the Dental Practitioner. Preprints, 2021.

Aoun G, El-Outa A, Nasseh I. Sialoliths: A Retrospective Radiological Study. Preprints, 2021.

Aoun G. Thyroid Dysfunction: Risk and Management in Dentistry. Preprints, 2021.

Aoun G. Management of Patients with Addison’s Disease in Dentistry: An Overview. Preprints, 2020.

Aragao M, Gomes F, de Melo L, Corona S. Brazilian dental students and COVID-19: a survey on knowledge and perceptions. medRxiv, 2020.

Aragao M, Gomes F, Coelho C, de Melo L, Corona S. Where do Brazilian dental students seek information about COVID-19? medRxiv, 2020.

Artzi Z, Sudri S, Platner O, Kozlovsky A. Regeneration of the Periodontal Apparatus in Aggressive Periodontitis Patients. Preprints, 2019.

Bachtiar B, Bachtiar E, Sunarto H, Soeroso Y, Sulijaya B, Theodorea C, Pratomo I, Yudhistira, Kusumaningrum A, Efendi D. ACE2 gene expression and inflammatory conditions in periodontal microenvironment of COVID-19 patients with and without diabetes evaluated by qPCR. medRxiv, 2022.

Baez V, Cruz M, Jurado L, Castro L, Nuñez F. Quality of Research in Residents of Medical Specialties after a Standardized Digital Training Program with Rubrics. Preprints, 2022.

Baima G, Citterio F, Romano F, Mariani G, Picollo F, Buduneli N, Aimetti M. Impact of surface decontamination and systemic antimicrobials for surgical treatment of peri-implantitis: A systematic review and meta-analysis of randomized clinical trials. medRxiv, 2021.

Bakhsh A, Alturkstani H, Alharbi R, Alrefai H, Badeeb T, Altouki N, Jamleh A. Interfacial Gap Assessment of Two Dental Adhesives and Polymer-Based Resin Composites Using Cross-Polarization Optical Coherence Tomography. Preprints, 2018.

Balasubramanian M, Hasan A, Ganbavale S, Alolayah A, Gallagher J. Planning the Future Oral Health Workforce: A Rapid Review of Supply, Demand and Need Models, Data Sources and Skill Mix Considerations. Preprints, 2021.

Banavar G, Ogundijo O, Julian C, Toma R, Camacho F, Torres P, Hu L, Kenny L, Vasani S, Batstone M. Detecting salivary host-microbiome RNA signature for aiding diagnosis of oral and throat cancer. medRxiv, 2022.

Barbato L, Gonnelli A, Baldi N, Francioni E, Tonelli P. Effect of Mechanical Stimulation on Clinical and Radiographical Healing in a Post-Extraction Site. Preprints, 2018.

Baridneh Y, Malek M, Mohamadian F, Amini S, Shamshirband S. Dental Scaling Instrument Selection by Applying Multi-Attribute Decision Making (MADM) Approach. Preprints, 2019.

Bates A, Bates D. Analysing air particle quantity in a dental primary care setting. medRxiv, 2020.

Behera S, Choudhury G, Mohapatra A, Panigrahi S. An evaluation of the fracture resistance of endodontically treated extracted teeth restored with Titanium, Carbon and Fibre posts: An in-vitro randomized control trial. medRxiv, 2020.

Behzadian F, Borjali A, Chizari M. Design evaluation of a dental implant used in the jawbone D1 – D4 zones. medRxiv, 2020.

Bencharit S, Clark W, Stoner L, Chiang G, Sulaiman T. Recent Advancements in CAD/CAM Same-Day Dentistry in Practice and Education. Preprints, 2021.

Benzian H, Niederman R. A Dental Response to the COVID-19 Pandemic – Safe Aerosol-Free Emergent (SAFE) Dentistry. Preprints, 2020.

Berman S, Sharp G, Lewis S, Blakey R, Davies A, Humphries K, Wren Y, Sandy J, Stergiakouli E. Prevalence and Factors Associated with Behavioural Problems in 5-year-old Children Born with Cleft Lip and/or Palate from the Cleft Collective. medRxiv, 2021.

Bhavasar R, Ajith N, Bhavasar R, Shah A, Vaswani V. Comparative evaluation of Oral lesions: “Tale - the Covid 19 Tells”. medRxiv, 2022.

Blume O, Hoffmann L, Donkiewicz P, Wenisch S, Back M, Schnettler R, Barbeck M. Treatment of Severely Resorbed Maxilla Due to Peri-Implantitis by Guided Bone Regeneration Using a Customized Allogenic Bone Block: A Case Report. Preprints, 2017.

Botelho J, Mascarenhas P, Mendes J, Machado V. Network Protein Interaction in Parkinson’s Disease and Periodontitis Interplay: A Bioinformatic Analysis. Preprints, 2020.

Botelho J, Machado V, Mascarenhas P, Alves R, Cavacas M, Mendes J. Fine-Tuning Multilevel Modeling of Risk Factors Associated with Nonsurgical Periodontal Treatment Outcome. Preprints, 2019.

Botelho J, Mascarenhas P, Viana J, Proença L, Orlandi M, Leira Y, Chambrone L, Mendes J, Machado V. An umbrella review of the evidence linking oral health and systemic health: from the prevalence to clinical and circulating markers. medRxiv, 2022.

Botelho J, Machado V, Leira Y, Proença L, Mendes J. Periodontal inflammation mediates the link between homocysteine and high blood pressure. medRxiv, 2021.

Bucur S, Moraru A, Adamovits B, Bud E, Olteanu C, Vaida L. Psychometric Properties of SCARED-C Scale in a Romanian Community Sample and its Future Utility for Dental Practice. Preprints, 2021.

Buffoli B, Grazetti G, Calza S, Scotti E, Borsani E, Cappa V, Rimondini L, Mensi M. Mensi. Preprints, 2018.

Burgard N, Kienitz M, Jourdan C, Rüttermann S. The Influence of Modified Experimental Dental Resin Composites on the Initial In Situ Biofilm – A Triple-Blinded Randomized Controlled Split-Mouth Trial. Preprints, 2021.

Carda-Diéguez M, Rosier B, Lloret S, Llena C, Mira A. The tongue biofilm metatranscriptome identifies metabolic pathways associated with halitosis and its prevention. medRxiv, 2021.

Carralero J, Pérez A, García A, M.J. M. Evolution of Necrotizing Periodontitis in a Patient with Multiple Sclerosis. Preprints, 2022.

Carter E, Currie C, Asuni A, Goldsmith R, Toon G, Horridge C, Simpson S, Donnell C, Greenwood M, Walton G, Cole B, Durham J, Holliday R. The first 6 weeks – setting up a UK urgent dental care centre during the COVID-19 pandemic, medRxiv, 2020.

Chandra R, Bhagyasree M, Sathwika P, Reddy A. Effect of two methods of soft tissue augmentation for socket closure on soft tissue landmarks and ridge dimensions-Results from software-based analysis of clinical data. medRxiv, 2022.

Chandra R, Shravan T. CORRELATION BETWEEN MEASURED PARAMETERS OF RISK AND PROGNOSIS IN SUBJECTS WITH CHRONIC PERIODONTITIS. medRxiv, 2022.

Chapain K, Rampal K, Pokhrel K, Adhikari C, Hamal D, Pokhrel K. Factors affecting oral health problems among school children in Kaski District, Nepal. medRxiv, 2022.

Chen C, Hsu K, Hsiao S, Tseng Y. The Correlation Among Gripping Volume, Insertion Torque, and Pullout Strength of Micro-implant. Preprints, 2020.

Choudhary S, Durkin M, Stoeckel D, Steinkamp H, Thornhill M, Lockhart P, Babcock H, Kwon J, Liang S, Biswas P. A Comparison of Aerosol Mitigation Strategies and Aerosol Persistence in Dental Environments. medRxiv, 2021.

Conte G, Pacino S, Urso T, Emma R, Caponnetto P, Cibella F, Polosa R. REPEATABILITY OF A DIGITAL IMAGING TECHNOLOGY FOR DENTAL PLAQUE QUANTITATION (QRAYCAM™ PRO) IN CURRENT, FORMER AND NEVER SMOKERS – STUDY PROTOCOL. medRxiv, 2021.

Conte G, Pacino S, Urso S, Emma R,Cibella F, Pedullà E, Polosa R. REPEATABILITY OF A CALIBRATED DIGITAL SPECTROPHOTOMETER FOR DENTAL SHADE EVALUATION IN CURRENT, FORMER AND NEVER SMOKERS – STUDY PROTOCOL. medRxiv, 2021.

Conte, et al. International Randomized Controlled Trial Evaluating Changes in Oral Health in Smokers after Switching to Combustion-Free Nicotine Delivery Systems: SMILE Study protocol. medRxiv, 2021.

Conway DI C. SARS-CoV-2 positivity in asymptomatic-screened dental patients. medRxiv, 2021.

Cosola S, Marcocini S, Boccuzzi M, Fabris G, Covani U, Peñarrocha-Diago M, Peñarrocha-Oltra D. Radiological Outcomes of Bone-Level and Tissue-Level Dental Implants: Systematic Review. Preprints, 2020.

Currie C, Palmer J, Stone S, Brocklehurst P, Aggarwal V, Dorman P, Pearce M, Durham J. Persistent Orofacial Pain Attendances at General Medical Practitioners. medRxiv, 2022.

Czumbel L, Kiss S, Farkas N, Mandel I, Hegyi A, Nagy A, Lohinai Z, Szakács Z, Hegyi P, Steward M. Saliva as a Candidate for COVID-19 Diagnostic Testing: A Meta-Analysis. medRxiv, 2020.

Darvell B. Bioactivity – Symphony or Cacophony?. Preprints, 2021.

DDS A, Zollanvari A. Modulating Cancer Progression from Leukoplakia via Bayesian Gene Networks. Preprints, 2021.

Devlin H, Williams T, Graham J, Ashley M. The ADEPT Study, A Comparative Study of Dentists’ Ability to Detect Enamel-only Proximal Caries in Bitewing Radiographs With and Without the use of AssistDent® Artificial Intelligence Software. medRxiv, 2021.

Devlin H, Ashley M, Williams T, Purvis B, Roudsari R. A Pilot Comparative Study of Dental Students’ Ability to Detect Enamel-only Proximal Caries in Bitewing Radiographs With and Without the use of AssistDent® Deep Learning Software. medRxiv, 2020.

Diachkova E, Popova S, Arazashvili L, Petruk P, Cherkesov I. Complicated Mandible Fracture Treatment With Xenogenic Bone Graft: A Case Report. Preprints, 2022.

Din A, Hindocha A, Patel T, Sudarshan S, Cagney N, Koched A, Mueller J, Seoudi N, Morgan C, Shahdad S, Fleming P. Quantitative particle analysis of particulate matter release during orthodontic procedures: A pilot study. medRxiv, 2020.

Dong Q, Kuria A, Weng Y, Liu Y, Cao Y. Impacts of the COVID-19 epidemic on the department of stomatology in a tertiary hospital: a case study in the General Hospital of the Central Theater Command, Wuhan, China. medRxiv, 2020.

Dot G, Schouman T, Chang S, Rafflenbeul F, Kerbrat A, Rouch P, Gajny L. Automatic Three-Dimensional Cephalometric Landmarking via Deep Learning. medRxiv, 2022.

Dot G, Rafflenbeul F, Kerbrat A, Rouch P, Gajny L, Schouman T. Three-dimensional cephalometric landmarking and Frankfort horizontal plane construction: reproducibility of conventional and novel landmarks. medRxiv, 2021.

Dot G, Schouman T, Dubois G, Rouch P, Gajny L. Fully automatic segmentation of craniomaxillofacial CT scans for computer-assisted orthognathic surgery planning using the nnU-Net framework. medRxiv, 2021.

Dudding, et al. A clinical observational analysis of aerosol emissions from dental procedures. medRxiv, 2021.

Ebrahimi T, Farokhi M. Effectiveness of Oral Health Education Interventions on Oral Health Literacy Levels in Adults; A Systematic Review. medRxiv, 2022.

El-Bialy T. The Effect of High Frequency Vibration on Tooth Movement and Alveolar Bone in Non-Growing Skeletal Class II High Angle Orthodontic Patients: Case Series. Preprints, 2020.

Elenora Martina B, Chandra R, Santosh V, Reddy G, Reddy A. A COMPARATIVE EVALUATION OF NON-INCISED PAPILLAE AND ENTIRE PAPILLA PRESERVATION SURGICAL APPROACHES IN TREATMENT OF INTRABONY DEFECTS – A CLINICAL AND RADIOGRAPHIC STUDY. medRxiv, 2022.

Fahim A, Rana S, Atif S, Basha S, Haider I, Alam M, Nagarajappa A. From Text to E-text: Perceptions of Medical, Dental and Allied Students About E-learning. Preprints, 2022.

Farnell D, Galloway J, Zurov A, Richmond S, Marshall D, Rosin P, Al-Meyah K, Pirttiniemi P, Lähdesmäki R. What’s in a Smile? Initial Analyses of Dynamic Changes in Facial Shape and Appearance. Preprints, 2018.

Feher B, Gruber R, Gahleitner A, Celar A, Necsea P, Ulm C, Kuchler U. Angular changes in implants placed in the anterior maxillae of adults: A cephalometric pilot study. medRxiv, 2020.

Felice G, Bianca D, Matteo N, Paolo C, Vinci R. Tilted Implants and Sinus Floor Elevation Techniques Compared in Posterior Edentulous Maxilla: Retrospective Clinical Study of Four Years Follow-up. Preprints, 2022.

Ferrés-Amat E, Madhoun A, Ferrés-Amat E, Carreo N, Barrajas M, Al-Madhoun A, Ferrés-Padró E, Marti C, Atari M. Comparison of 0.12% Chlorhexidine and a New Bone Bioactive Liquid, BBL, Mouthrinses on Oral Wound Healing: A Randomized, Double Blind Clinical Human Trial. Preprints, 2022.

Fornaini C, Merigo E, Poli F, Rocca J, Selleri S, Lagori G, Cucinotta A. Dentine Laser Welding: A New Help for Fractured Teeth? A Preliminary Ex Vivo Study. Preprints, 2018.

Fountoulaki G, Thurzo A. Change in the Constricted Airway in Patients after Clear Aligner Treatment: Retrospective Study. Preprints, 2022.

Fu L, Ling C, Jin Z, Luo J, Palma-Chavez J, Wu Z, Zhou J, Zhou J, Donovan B, Qi B. A more compact photoacoustic imaging system to detect periodontitis. medRxiv, 2021.

Fu Y, Xu H. Expression of deubiquitinases in human gingiva and cultured human gingival fibroblasts. medRxiv, 2021.

Garcia B, Acosta N, Tomar S, Roesch L, Lemos J, Mugayar L, Abranches J.Association of Streptococcus mutans harboring bona-fide collagen binding proteins and Candida albicans with early childhood caries recurrence. medRxiv, 2021.

Gebretsadik H. An update on oral clinical courses among patients with severe acute respiratory syndrome coronavirus 2 (SARS-CoV-2) infection: A clinical follow-up (a prospective prevalent cohort) study. medRxiv, 2022.

Germano F, Testi D, Campagnolo L, Scimeca M, Arcuri C. Cell-wall-deficient Bacteria in Oral Biofilm: Association with Periodontitis. medRxiv, 2020.

Ghalandari M, Malek M, Alizadeh H, Ghalandari F, Mosavi A, Shamshirband S, Chau K. Effect of Needle Size and Volumetric Flow Rate on Root Canal Irrigation: A Numerical Investigation. Preprints, 2019.

Gkekas A, Varenne B, Benzian H, Stauf N, Listl S. Affordability of Essential Medicines: The Case of Fluoride Toothpaste in 78 Countries. Preprints, 2022.

Goldrick N, O’Keefe E. Surveillance of COVID-19 cases associated with dental settings using routine health data from the East of Scotland with a description of efforts to break chains of transmission from October 2020 to December 2021. medRxiv, 2022.

Gormley A, Haworth S, Simancas-Pallares M, Holgerson P, Esberg A, Shrestha P, Divaris K, Johansson I. Subtypes of early childhood caries predict future caries experience. medRxiv, 2022.

Grande M, Teixeira M, Pelegrine A, Lopes G, Campos J, Nishioka R. Effect of Maxillary Implants Region and Loading Condition in the Stress Distribution of Implant-Supported Full-Arch Prosthesis: 3D-FEA. Preprints, 2021.

Grimm W, Didenko5 N, Dhingra K, Dolgalev A, Enukashvily N, Fritsch T, Giesenhagen5 B, Ivolgin D, Vukovic M. Isolation of Neural Crest-derived Stem Cells for Therapeutic Use in Regenerative Periodontology. Preprints, 2020.

Grzech-Leśniak K, Bencharit S, Skrjanc L, Kanduti D, Matys J, Deeb J. Utilization of Er:YAG Laser in Retrieving and Reusing of Lithium Disilicate and Zirconia Monolithic Crowns in Natural Teeth: An in Vitro Study. Preprints, 2020.

Gurr A, Higgins D, Henneberg M, Kumaratilake J, Brook O’Donnell M, McKinnon M, Brook A. Large Volume Micro-CT scanning of the dentoalveolar complex as a tool to evaluate dental health in an archaeological sample compared with traditional methods. medRxiv, 2022.

Hadilou M, Dolatabadi A, Ghojazadeh M, Hosseinifard H, Alizadeh Oskuee P, Pournaghi Azar F. Which surface treatment improves the long-term repair bond strength of aged methacrylate-based composite resin restorations? A systematic review and network meta-analysis. medRxiv, 2022.

Han H, Morse Z, Koziol-McLain J, Lees A. Oral health professionals and child abuse and neglect: a scoping review protocol. medRxiv, 2022.

Hernández-Olivos R, Muñoz M, Núñez E, Camargo-Ayala P, Garcia-Huidobro J, Pereira A, Nachtigall F, Santos L, Rivera C. Salivary proteome of aphthous stomatitis reveals the participation of vitamin metabolism, nutrients, and bacteria. medRxiv, 2021.

Hernández-Vásquez A, Barrenechea-Pulache A, Comandé D, Azañedo D. Mouth-rinses and SARS-CoV-2 viral load in saliva: A living systematic review. medRxiv, 2021.

Hobson R, Pabary S, Alamani K, Badminton K. Aerosols created by dental procedures in a primary care setting. medRxiv, 2020.

Homolak J, Tomljanovic D, Milosevic M, Vrazic D, Zivkovic M, Budimir I, Nikolic B, Muslim A, Ljubicic N, Nikolic M. A Cross-Sectional Study of Hepatitis B and Hepatitis C Knowledge Among Dental Medicine Students at the University of Zagreb. Preprints, 2020.

Huang Q, Rasubala L, Gracely R, Khan J, Eliav E, Ren Y. Pain management after dental extractions – non-opioid combination analgesics minimize opioid use for acute dental pain. medRxiv, 2022.

Huang Y, Lee S. Deep Learning for Caries Detection using Optical Coherence Tomography. medRxiv, 2021.

Huang Q, Marzouk T, Cirligeanu R, Malmstrom H, Eliav E, Ren Y. Ventilation rate assessment by carbon dioxide levels in dental treatment rooms. medRxiv, 2021.

Iandolo A, Dagna A, Beltrami R, Poggio C, Malvano M, Amato A, Abdellatif D. Evaluation of the Actual Chlorine Concentration and the Required Time for Pulp Dissolution Using Different Naocl Irrigating Solutions. Preprints, 2018.

Ibrahim A, Alqalshy E, Abdel-Hafiz A, El-Rahman K, Alazzazi M, Omar M. Immunohistochemical Evaluation of CD31 and D2-40, Expression in Oral Squamous Cell Carcinoma. Preprints, 2022.

Ibrahim A, Elqalshy E, El-Mohamadi A, El-Rahman K, Alazzazi M.Roles of Proliferation and Angiogenesis in Locally Aggressive Biologic Behavior of Ameloblastoma versus Ameloblastic Fibroma. Preprints, 2022.

Innes N, Johnson I, Al-Yaseen W, Harris R, Jones R, Kc S, McGregor S, Robertson M, Wade W, Gallagher J. A Systematic Review of Droplet and Aerosol Generation in Dentistry. medRxiv, 2020.

Inquimbert C, Clement C, Couatarmanach A, Tramini P, Bourgeois D, Carrouel F. Oral Hygiene Practices and Knowledge among Adolescents Aged between 15 and 17 Years Old during Fixed Orthodontic Treatment: Multicentre Study Conducted in France. Preprints, 2022.

Jamal M, Shah M, Almarzooqi S, Aber H, Khawaja S, Abed R, Alkhatib Z, Samaranayake

L. Overview of Trans-National Recommendations for COVID-19 Transmission Control in Dental Care Settings. Preprints, 2020.

Jiang N, Grytten J, Kinge J. Inequality in access to dental services in a market-based dental care system. A population study from Norway 1975-2018. medRxiv, 2021.

Jin F, Song J, Luo Y, Wang B, Ding M, Hu J, Chen Z. Association between skull bone mineral density and periodontitis: evidence from the National Health and Nutrition Examination Survey (2011-2014). medRxiv, 2022.

Jung O, Becker J, Smeets R, Gosau M, Becker G, Kahl-Nieke B, Jung A, Proff P, Kopp A, Barbeck M, Köhne T. Surface Characteristics of Aesthetic Nickel-titanium and Beta-titanium Orthodontic Archwires Produced by Plasma Electrolytic Oxidation (PEO) - Primary Results. Preprints, 2019.

Jung O, Smeets R, Hartjen P, Schnettler R, Feyerabend F, Klein M, Walther F, Stangier D, Henningsen A, Rendenbach C, Heiland M, Barbeck B, Kopp A. Improved in Vitro Test Procedure for Full Assessment of the Cytocompatibility of Degradable Magnesium Based on ISO 10993-5/-12. Preprints, 2018.

Junior J, Biguetti C, Matsumoto M, Kudo G, Silva R, Saraiva P, Fakhouri W. Can Genetic Factors Compromise the Success of Dental Implants? A Systematic Review and Meta-Analysis. Preprints, 2018.

Kabil N, Eltaweil S. Reshuffling the Risk Factors of Severe Early Childhood Caries. Preprints, 2016.

Kalimuthu S, Cheung B, Yau J, Shanmugam K, Solomon A, Neelakantan P. A Novel Small Molecule, 1,3-di-m-tolyl-urea, Inhibits and Disrupts Multispecies Oral Biofilms. Preprints, 2020.

Kalman L, Piva A, Queiroz T, Tribst J. Biomechanical Behavior Evaluation of a Novel Hybrid Occlusal Splint-mouthguard for Contact Sports: 3D-FEA. Preprints, 2021.

Kang M, Neto H, Pelegrine A, Turssi C, Clemente-Napimoga J, Napimoga M. Survival rate of dental implants installed by residents attending an implantology Program in Brazil: a 52-month retrospective analysis. medRxiv, 2022.

Katsiroumpa A, Galanis P, Diamantis I, Georgikopoulou S, Katsoulas T, Katsimperi E, Konstantinou E. Impact of diabetes mellitus on the stabilization and osseointegration of dental implants: a systematic review. medRxiv, 2022.

Katsiroumpa A, Galanis P, Diamantis I, Georgikopoulou S, Katsoulas T, Konstantakopoulou O, Katsimperi E, Konstantinou E. Impact of diabetes mellitus on the stabilization and osseointegration of dental implants: a retrospective study in Greece. medRxiv, 2022.

Keeper J, Skaret L, Thakkar-Samtani M, Heaton L, Sutherland C, Vela K, Amaechi B, Jablonski-Momeni A,Young D, MacLean J. Systematic Review and Meta-Analysis on the Effect of Self-Assembling Peptide P11-4 on Initial Caries Lesions. medRxiv, 2022.

Kelly A, Kaliskova A, Küchler E, Romanos H, Lips A, Costa M, Modesto A, Vieira A. Measuring the Microscopic Structures of Human Dental Enamel Can Predict Caries Experience. Preprints, 2020.

Khalil G, Aoun G, Khalifeh B, Zeinoun T. Segmental Mandibular Resection for Conventional Ameloblastoma. Preprints, 2021.

Kiliç B, Önal-Süzek T. Aksoy S. Comparative Analysis of three machine learning models for Early Prediction of Skeletal Class-III Malocclusion from Profile Photos. medRxiv, 2022.

Kinariwala N, Samaranayake L, Perera I, Patel Z. Knowledge, Awareness and Perceptions of Coronavirus Disease 2019 (COVID-19) in a Cohort of Indian Dental Professionals: A Questionnaire-Based Study. Preprints, 2020.

Kolahi J, Khazaei S, Iranmanesh P. Science Map of the Highly Tweeted Endodontic Articles. medRxiv, 2021.

Kori J, Saleem F, Ullah S, Azim M. Characterization of Oral bacteriome dysbiosis in type 2 diabetic patients. medRxiv, 2020.

Kumar S, Saxena S, Atri M, Chamola S. Effectiveness of the Covid-19 vaccines in preventing infection in dental practitioners – results of a cross-sectional ‘questionnaire-based’ survey. medRxiv, 2021.

Kumar R, Karumaran S, Kattula D, Thavarajah R, Anusa A. Perceived Stress and Psychological (Dis)Stress among Indian Endodontists During COVID19 Pandemic Lock down. medRxiv, 2020.

Lai S, Novillo F, Cárdenas G, Verdugo F, Rada G. Hyaluronic acid for tissue and bone regeneration after tooth extraction. Systematic review and meta-analysis. medRxiv, 2020.

Lee Y, Auh Q, An J, Kim T. Effects of Clinical Characteristics on Sleep Quality in Patients with Chronic Temporomandibular Disorders. Preprints, 2021.

Lee J, Lee J, Song H, Son M, Li L, Rhyu I, Lee Y, Koo K, An J, Kim J. Diagnostic Models for Screening of Chronic Periodontitis with Cytokines and Microbiologic Profiles in Saliva. Preprints, 2020.

Leiva-Sabadini C, Schuh C, Barrera N, Aguayo S. Ultrastructural characterisation of young and aged dental enamel by atomic force microscopy. medRxiv, 2022.

LI D, LEUNG Y. Disorders: Current Concepts and Controversies in Diagnosis and Management. Preprints, 2021.

Li D, Wong N, Li S, McGrath C, Leung Y. Timing of Arthrocentesis in the Management of Temporomandibular Disorders:A Systematic Review and Meta-analysis. Preprints, 2020.

Li X, Liu D, Sun Y, Yang J, Yu Y. Association of genetic variants in enamel-formation genes with dental caries: A meta- and gene-cluster analysis. medRxiv, 2020.

Liang Y, Huan J, Li J, Jiang C, Fang C, Liu Y. Recovering Mandibular Morphology after Disease with Artificial Intelligence. medRxiv, 2020.

Lim Q, Taylor K, Dudding T. The effects of modifiable maternal pregnancy exposures on offspring molar-incisor hypomineralisation: A negative control study. medRxiv, 2022.

Limiroli E, Calò A, Limiroli A, Rasperini G. Radiographic Ratios for Classifying Furcation Anatomy. Proposal of a New Evaluation Method and an Intra-Rater and Inter-Rater Operator Reliability Study. Preprints, 2022.

Liu Z, Zhang T, Wu K, Li Z, Chen X, Jiang S, Du L, Lu S, Lin C, Wu J. Metagenomic Analysis Reveals A Possible Association Between Respiratory Infection and Periodontitis. medRxiv, 2020.

Llandro H, Allison J, Currie C, Edwards D, Bowes C, Durham J, Jakubovics N, Rostami N, Holliday R. Evaluating splatter and settled aerosol during orthodontic debonding: implications for the COVID-19 pandemic. medRxiv, 2020.

Lulu P, Nanyingi M. Factors influencing adoption of oral health promotion by antenatal care providers in Moyo district, North-Western Uganda. medRxiv, 2022.

Luzzi V, Ierardo G, Bossù M, Polimeni A. COVID-19: Pediatric Oral Health During and After the Pandemics. Preprints, 2020.

Machado V, Botelho J, Mascarenhas P, Mendes J, Delgado A. A Systematic Review and Meta-Analysis on Bolton’s Ratios: Normal Occlusion and Malocclusion. Preprints, 2019.

Machado V, Botelho J, Neves J, Proença L, Alves R, Cavacas M, Delgado A, Mendes J. The Prevalence of Periodontal Diseases in Portugal and Correspondent Digital National Awareness: Analysis of Data from Global Burden of Disease, Directorate-General of Health and Google Trends for the Period 2004-2017. Preprints, 2019.

Macrì M, Toniato E, Murmura G, Varvara G, Festa F. Midpalatal Suture Density CBCT Evaluation, Sex Gender and Growth Pattern Related Variability in 392 Adolescents Treated With Rapid Maxillary Expander Appliance. Preprints, 2022.

Mahajan A, Asi K, Kaushal A, Bhatia V, Mahajan P. Evidence Based Decision Making in Dental Treatment During COVID-19 Outbreak. Preprints, 2020.

Maida C, Marcus M, Xiong D, Ortega-Verdugo P, Agredano E, Huang Y, Zhou L, Lee S, Shen L, Hays R, Crall J, Liu H. Perceptions of Teachers and School Nurses on Child and Adolescent Oral Health. Preprints, 2022.

Maria Ferreira A, Eveloff R, Freire M, Rodrigues Santos M. Clinical and Inflammatory Factors Influencing Constipation and Quality of Life in Cerebral Palsy. medRxiv, 2019.

Martínez-Martínez E, Medina-Solís C, Alpuche J. Total Edentulism and Its Epidemiological Surveillance in Oaxaca, Mexico from 2009–2019. Preprints, 2021.

Massignan C, Soares J, de Souza Pires M, Dick B, Porporatti A, De Luca Canto G, Bolan M. Parental acceptance toward behavior guidance techniques for pediatric dental visits: a meta-analysis. medRxiv, 2020.

Matarese G, Matarese M, Picciolo G, Fiorillo L, Isola G. Evaluation of Low-Level Laser Therapy with Diode Laser for the Enhancement of the Orthodontic Tooth Movement: a Split-Mouth Study. Preprints, 2018.

Medina C, Ueda H, Kunimatsu R, Tanimoto K. Changes in the position of the hyoid bone in skeletal Class II children post-functional Activator therapy. medRxiv, 2020.

MENSI M, COCHIS A, SORDILLO A, UBERTI F, RIMONDINI L. Biofilm removal and bacterial re-colonization inhibition of a novel erythritol/chlorhexidine air-polishing powder on titanium disks. Preprints, 2018.

Moore C, Law J, Pham C, Joe Chang K, Chen C, Jokerst J. High-resolution ultrasonography of gingival biomarkers for periodontal diagnosis in healthy and diseased subjects. medRxiv, 2021.

Moraes R, Correa M, Daneris A, Queiroz A, Lopes J, Lima G, Cenci M, D’Avila O, Pannuti C, Pereira-Cenci T, Demarco F. Email vs. Instagram recruitment strategies for online survey research. medRxiv, 2020.

Moraes R, Correa M, Queiroz A, Daneris A, Lopes J, Pereira-Cenci T, D’Avila O, Cenci M, Lima G, Demarco F. COVID-19 challenges to dentistry in the new pandemic epicenter: Brazil. medRxiv, 2020.

Mozaffari H, Zavattaro E, Abdolahnejad A, Jornet P, Omidpanah N, Sharifi R, Sadeghi M, Shooribi M, Safaei M. Serum and Salivary IgA, IgG, and IgM Levels in Oral Lichen Planus: A Systematic Review and Meta-Analysis of Case-Control Studies. Preprints, 2018.

Murererehe J, Malele-Kolisa Y, Niragire F, Yengopal V. Prevalence of dental caries and associated risk factors among HIV-positive and HIV-negative adults at an HIV clinic in Kigali, Rwanda. medRxiv, 2022.

Nagarajan R, Panny A, Ryan M, Murphy S, Vujicic M, Nycz G. Forecasting Preventive Dental Quality Measures. medRxiv, 2021.

Nakano L, Gomes L, Queiroz T, Paes-Junior T. Effect of Abutment Type and Tightening Sequence on Torque Maintenance Capacity after Mechanical Cycling in Splinted Implant-Supported Restorations. Preprints, 2021.

Neupaul P, Mahomed O. Influence of Parents’ Oral Health Knowledge and Attitudes on Oral Health Practices of Children (5-12 Years) in a Rural School in KwaZulu Natal, South Africa: A Cross Sectional Survey. Preprints, 2020.

Nicozisis J, Brigham G, Sparaga J, Shipley T. Effects of a 120Hz High-Frequency Acceleration Device on Orthodontic Discomfort. Preprints, 2018.

Nikinmaa S, Moilanen N, Sorsa T, Rantala J, Alapulli H, Kotiranta A, Auvinen P, Kankuri E, Meurman J, Pätilä T. Small-form-factor LED-based light application surface for indocyanine green-mediated antibacterial photothermal therapy. medRxiv, 2020.

Niraula N, Acharya R, Humagain M, Khurshid Z, Adanir N, Rokaya D. Dental-Facial Midline: An Esthetic Based Classification. medRxiv, 2021.

Nishimoto S, Saito T, Ishise H, Fujiwara T, Kawai K, Kakibuchi M. Machine learning-based noise reduction for craniofacial bone segmentation in CT images. medRxiv, 2022.

Nishimoto S, Kawai K, Fujiwara T, Ishise H, Kakibuchi M. Locating cephalometric landmarks with multi-phase deep learning. medRxiv, 2020.

Nørskov-Lauritsen N, Claesson R, Jensen A, Höglund-Åberg C, Haubek D. Aggregatibacter Actinomycetemcomitans: Clinical Significance of a Pathobiont Subjected to Ample Changes in Classification and Nomenclature. Preprints, 2019.

Novillo F, Lai S, Cárdenas G, Verdugo F, Rada G. Periodontal therapy on disease activity of Rheumatoid Arthritis. Systematic review and meta-analysis. medRxiv, 2020.

Nulty A, Lefkaditis C, Zachrisson P, Tonder Q, Yar R. A Clinical Study Measuring Dental Aerosols with and without a HVE Device. Preprints, 2020.

Nulty A. A Review of Evidence: Using Respirators to prevent Sars-Cov-2 Transmission. Preprints, 2020.

Okawa H, Kondo T, Hokugo A, Cherian P, Campagna J, Lentini N, Sun S, Sung E, Chiang S, Lin Y, Ebetino F, John V, McKenna C, Nishimura I. Mechanism of bisphosphonate-related osteonecrosis of the jaw (BRONJ) revealed by targeted removal of legacy bisphosphonate from jawbone using equilibrium competing inert hydroxymethylene diphosphonate. medRxiv, 2021.

Okuji D, Odusanwo O, Wu Y, Yeh S, Dhar S. Sucrose-stimulated Salivary pH as an Adjunct to Caries Risk Assessment. medRxiv, 2022.

Okuji D, Jivraj A, Wu Y. From Volume to Value: Comparative value, quality, and cost between three treatment modalities for early childhood caries. medRxiv, 2022.

Okunev I, Frantsve-Hawley J, Tranby E. Trends in National Opioid Prescribing for Dental Procedures among Medicaid Patients. medRxiv, 2021.

Okunseri C, Frantsve-Hawley J, Thakkar-Samtani M, Okunev I, Tranby E. Estimation of Oral Disease Burden from Claims and Self-Reported Data. medRxiv, 2021.

Oliveira L, Borges T, Soares R, Buzzi M, Kuckelhaus S. Methodological Variations Affect the Release of VEGF in Vitro and Fibrinolysis’ Time from Platelet Concentrates. Preprints, 2020.

Omura T, Matsuyama M, Nishioka S, Sagawa S, Seto M, Naoe M. Association Between the Swallowing Reflex and the Development of Aspiration Pneumonia in Patients with Dysphagia Admitted to Long-term Care Wards -A Prospective Cohort Study of 60 Days-. medRxiv, 2020.

Ou Q, Placucci R, Danielson J, Anderson G, Olin P, Jardine P, Madden J, Yuan Q, Grafe T, Shao S, Hong J, Pui D. Characterization and mitigation of aerosols and splatters from ultrasonic scalers. medRxiv, 2021.

Pakarinen S, Saarela R, Välimaa H, Heikkinen A, Kankuri E, Noponen M, Alapulli H, Tervahartiala T, Räisänen I, Sorsa T, Pätilä T. A Randomized Trial of Home-Applied Dual-Light Photodynamic Therapy in Stable Chronic Periodontitis-Three-Month Interim Results. Preprints, 2022.

Pan J, Li N, Tang Q, Li G, Hou Y, Wang L, Yu W. Gelatin Nanoparticles Loaded with PMX-53 Prevents Alveolar Bone Loss in Miniature Swines Model of Periodontitis. Preprints, 2017.

Panta P, Felix D, Andreadis D, Seshadri M. Linear Oral Lichen Planus: A Rare Presentation. Preprints, 2020.

Papadaki S, Douglas G, HaniBani A, Kang J, Gender Differences in Caries and Periodontal Status in UK Children. medRxiv, 2021.

Paraizo M, Botelho J, Machado V, Mendes J, Alves R, Mascarenhas P, Cardoso J. Dental Implant Failure Rate and Marginal Bone Loss in Transplanted Patients: A Systematic Review and Meta-Analysis. Preprints, 2020.

Pardal-Peláez B, Pardal-Refoyo J, Montero-Martín J, González-Serrano J, López-Quiles-Martínez J. Classification of sinonasal pathology associated with dental pathology or dental treatment. medRxiv, 2020.

Park S, Lee J. Effects of Type 2 Diabetes Mellitus on Osteoclast Differentiation, Activity, and Cortical Bone Formation in Postmenopausal MRONJ Patient. Preprints, 2022.

Passarelli P, D’Addona A, Ferro L, Lopez M. Management of Patients with Coagulation Disorders Undergoing Minor Oral Surgery. Preprints, 2022.

Paul B, Sierra M, Xu F, Crystal Y, Li X, Saxena D, Ruff R. Microbial Population Shift and Metabolic Characterization of Silver Diamine Fluoride Treatment Failure on Dental Caries. medRxiv, 2020.

Pereira L, Mourão C, Alves A, Resende R, Uzeda M, Louro R, Calasans-Maia M. Standardized in vivo Evaluation of Biocompatibility and Bioresorption of a New Synthetic Membrane for Guided Bone Regeneration. Preprints, 2019.

Pereira A, Brito G, Lima M, Júnior A, Silva E, Rezende A, Bortolin R, Galvan M, Pirih F, Júnior R, Medeiros C, Guerra G, Araújo A. Metformin Hydrochloride-Plga Nanoparticles in Diabetic Rats in A Periodontal Disease Experimental Model. Preprints, 2018.

Poluha R, Rizzatti-Barbos C, Pinzón N, Silva B, Almeida A, Ernberg M, Manso A, Bonjardim L, Canales G. Does Botulinum Toxin Type–a Improve Mandibular Motion and Muscle Sensibility in Myofascial Pain Tmd Subjects? A Randomized, Controlled Clinical Trial. Preprints, 2022.

Proctor D, Seiler C, Burns A, Walker S, Jung T, Weng J, Sastiel S, Rajendran Y, Kapila Y, Millman M. Spatial patterns of dental disease in patients with low salivary flow. medRxiv, 2021.

Qing M, Shang Q, Yang D, Peng J, Deng J, Jiang L, Li J, Zhou Y, Xu H, Chen Q. CD8+ tissue-resident memory T cells triggered the erosion of oral lichen planus by the cytokine network. medRxiv, 2022.

Reed S, Fan S, Wagner C, Lawson A. Predictors of Developmental Defects of Enamel in the Primary Maxillary Central Incisors using Bayesian Model Selection. medRxiv, 2022.

Relvas M, Regueira-Iglesias A, Balsa-Castro C, Salazar F, Pacheco J, Cabral C, Henriques C, Tomás I. Assessing the impact of dental and periodontal statuses on the salivary microbiome: a global oral health scale. medRxiv, 2020.

Riad A, Issa J, Chuchmova V, Slezakova S, Gomaa E, Pokorna A, Klugarova J, Klugar M. Oral ulcers of COVID-19 patients: a scoping review protocol. medRxiv, 2021.

Ribas Pérez D, Olivera R, Mendoza-Mendoza A. Knowledge and aptitude of early childhood, primary and/or secondary education teachers referred to first aid measures in dental trauma in the province of Seville (Spain.). medRxiv, 2022.

Ribas P, Olivera R, Mendoza-Mendoza A. Knowledge and aptitude of early childhood, primary and/or secondary education teachers referred to first aid measures in dental trauma in the province of Seville (Spain.). medRxiv, 2022.

Rivera C, Muñoz-Pastén M, Muñoz-Núñez E, Hernández-Olivos R. Recurrent aphthous stomatitis affects quality of life. medRxiv, 2022.

Romandini M, Lima C, Pedrinaci I, Araoz A, Soldini M, Sanz M. Prevalence and risk/protective indicators of peri-implant diseases: a university-representative cross-sectional study. medRxiv, 2020.

Romandini M, Lima C, Pedrinaci I, Araoz A, Soldini M, Sanz M. Clinical signs, symptoms, perceptions and impact on quality of life of patients suffering peri-implant diseases: a university-representative cross-sectional study. medRxiv, 2020.

Rovera A, Banducci L. A Novel Approach for Environmental Management of Dental Practice: The “Plan-Do-Check-Act” Model. Preprints, 2022.

Ruff R, Barry-Godin T, Niederman R. Non-inferiority of essential medicines for caries arrest and prevention in a school-based program: Results from the CariedAway pragmatic clinical trial. medRxiv, 2022.

Ruff R, Barry-Godin T, Whittemore R, Small T, Santiago-Galvin N, Sharma P. Severity of dental caries in New York City children receiving school-based prevention and the role of SARS-CoV-2: Results from the CariedAway pragmatic trial. medRxiv, 2022.

Ruff R, Barry-Godin T, Small T, Niederman R. Silver diamine fluoride, atraumatic restorations, and oral health-related quality of life -Results from the CariedAway cluster randomized trial. medRxiv, 2021.

Ruff R, Paul B, Sierra M, Xu F, Crystal Y, Li X, Saxena D. Treatment nonresponse in children receiving silver diamine fluoride for caries: A pilot study. medRxiv, 2020.

Salam A, Khan F. Periodontopathogens in oral cancer: a meta-analysis of bacterial taxa of the oral microbiome associated with risk factors of oral squamous cell carcinoma. medRxiv, 2022.

Salas G, Lai S, Verdugo-Paiva F, Requena R. Platelet-rich fibrin in third molar surgery. Systematic review and meta-analysis protocol. medRxiv, 2020.

Saleem K, Ahmad P, Karobari M, Kamal M, Asif J, Noorani T. Non-Dental Drugs A Dentist Should Know: A Review. Preprints, 2020.

Saleem F, Mujtaba G, Kori J, Hassan A, Azim M. Microbiome evaluation revealed salivary dysbiosis in addicts of betel nut preparations. medRxiv, 2020.

Samaranayake L, Fakhruddin K, Buranawat B, Panduwawala C. The Efficacy of Bio-aerosol Reducing Procedures Used in Dentistry: A Systematic Review. Preprints, 2020.

Sanchez-Perez A, Rosa-Vela T, Mateos-Moreno B, Jornet-Garcia A, Navarro-Cuellar C. Systematic Review and Meta-Analysis of the Use of Hyaluronic Acid Injections to Restore Interproximal Papillae. Preprints, 2021.

Sathwika P, Chandra R. Marginal Bone Loss and Aesthetics Around Zirconia and Titanium Dental Implants-A Metanalysis. medRxiv, 2020.

Scarano A, Lorusso F, Merla A, D’Arcangelo C, Oliveira P. Lateral Sinus Floor Elevation Performed with Trapezoidal and Triangular Flap Designs: Post Operative Pain Measurement through Thermal Infrared Imaging. A Randomized Pilot Study. Preprints, 2018.

Scardina G, Casella S, Bilello G, Messina P. Photobiomodulation Therapy in the Management of Burning Mouth Syndrome: Morphological Variations in the Capillary Bed. Preprints, 2020.

Schermann H, Schiffmann N, Ankory R, Shlaifer A, Yavnai N, Yoffe V, Natapov L. Methylphenidate use and restorative treatment in 13,000 young adults with ADHD. medRxiv, 2020.

Schneider C, Zitzmann N, Zemp E. Changes in dental care behaviour between 2002 and 2012 and its association with complete dentition in men and women in Switzerland. medRxiv, 2019.

Schuh C, Leiva-Sabadini C, Huang S, Barrera N, Bozec L, Aguayo S. Nanomechanical and molecular characterization of aging in dentinal collagen. medRxiv, 2021.

Schuller-Götzburg P, Forte T, Pomwenger W, Petutschnigg A, Watzinger F, Entacher K. Three-Dimensional Finite Element Analysis of Maxillary Sinus Floor Augmentation with Optimal Positioning of a Bone Graft Block. 2018.

Sellan P, Campaner L, Tribst J, Piva A, Andrade G, Borges A, Bresciani E, Ausiello P. Functional or Nonfunctional Cusps Preservation for Posterior Onlays in Indirect Composite or Glass-Ceramic: 3D FEA Study. Preprints, 2021.

Semprini J. Estimating the within-person change in dental service access measures during the COVID-19 Pandemic. medRxiv, 2022.

Seneviratne C, Balan P, Ki K, Udawatte N, Lai D, Lin D, Venkatachalam I, Sit J, Lin L, Oon L, Tin G, Ying J. Efficacy of commercial mouth-rinses on SARS-CoV-2 viral load in saliva: Randomized Control Trial in Singapore. medRxiv, 2020.

Shafiee S, Sofi-Mahmudi A, Behnaz M, Safiaghdam H, Sadr S. Iranian Dental Students and Specialists’ Knowledge and Attitude about Obstructive Sleep Apnea. medRxiv, 2020.

Shahdad S, Hindocha A, Patel T, Cagney N, Mueller J, Kochid A, Seoudi N, Morgan C, Fleming P, Din A. Fallow time determination in dentistry using aerosol measurement. medRxiv, 2021.

Sharifonnasabi F, Jhanjhi N, John J, Obeidy P, Shamshirband S, Rokny H, Baz M. Hybrid HCNN-KNN Model Enhances Age Estimation Accuracy in Orthopantomography. Preprints, 2022.

Shelke G, Marwaha R, Shah P, Challa S. Role of Patient’s Ethnicity in Seeking Preventive Dental Services at the Community Health Centers of South-Central Texas: A Cross-Sectional Study. Preprints, 2022.

Shipley T. High Frequency Acceleration Device Effect on Accelerated Aligner Exchange -A Pilot Study. Preprints, 2018.

Shirakawa H, Sakata K, Hato H, Sato J, Ohga N, Watanabe H, Iori T, Yamazaki Y, Kitagawa

Y. Ethyl Loflazepate as a Treatment for Patients with Idiopathic and Psychogenic Taste Disorder. Preprints, 2021.

Shoaee S, Shoaee F, Parsa P, Sofi-Mahmudi A. Dental caries among the elderly in Iran: a meta-analysis. medRxiv, 2020.

Sieger D, Korzinskas T, Jung O, Stojanovic S, Wenisch S, Smeets R, Gosau M, Schnetter R, Najman S, Barbeck M. The Addition of High Doses of Hyaluronic Acid to a Biphasic Bone Substitute Decreases the Proinflammatory Tissue Response. Preprints, 2019.

Simpson S, Sumner O, Holliday R, Currie C, Hind V, Lush N, Burbridge L, Cole B. Paediatric Dentistry and the coronavirus (COVID-19) response in the North East of England and North Cumbria. medRxiv, 2020.

Simpson, et al. A Novel de novo TP63 Mutation in Whole Exome Sequencing of a Syrian Family with Oral Cleft and Ectrodactyly. medRxiv, 2022.

Sneha K, Ajmera J, Chandra R. THE G-FORCE CONUNDRUM IN PRF GENERATION-MANAGEMENT OF A PROBLEM HIDDEN IN PLAIN SIGHT. medRxiv, 2020.

So M, Ellenikiotis Y, Husby H, Paz C, Seymour B, Sokal-Gutierrez K. Early Childhood Dental Caries, Mouth Pain, and Malnutrition in the Ecuadorian Amazon Region. Preprints, 2017.

Sofi-Mahmudi A, Iranparvar P, Shakiba M, Shamsoddin E, Mohammad-Rahimi H, Naseri S, Motie P, Mesgarpour B. Quality assessment of studies included in Cochrane oral health systematic reviews. medRxiv, 2020.

Sofi-Mahmudi A, Shamsoddin E, Ghasemi P, Nasser M, Mesgarpour B. The association between COVID-19-imposed lockdowns and online searches for toothache using Google Trends. medRxiv, 2020.

Sofi-Mahmudi A, Shamsoddin E, Ghasemi P, Bahar A, Azad M, Sadeghi G. Association of COVID-19-imposed lockdowns and online searches for toothache in Iran. medRxiv, 2020.

Sousa F, Machado V, Botelho J, Proença L, Mendes J, Alves R. Effect of A-PRF Application on Palatal Wound Healing after Free Gingival Graft Harvesting: A Prospective Randomized Study. Preprints, 2019.

Spivack E. Dental Care of the Homebound Patient with Myalgic Encephalomyelitis/chronic Fatigue Syndrome. Preprints, 2020.

Steiner G. After Mineralization, Mineralized Freeze-Dried Bone Allograft Particles are Exfoliated but not Resorbed. Preprints, 2019.

Steiner G.How Cadaver Bone Transplants Mineralize and Sclerotic Bone Fails. Preprints, 2018.

Stephen A, Dhadwal N, Nagala V, Gonzales-Marin C, Gillam D, Bradshaw D, Burnett G, Allaker R. Interdental and subgingival microbiota may affect the tongue microbial ecology and oral malodour in health, gingivitis and periodontitis. medRxiv, 2021.

Su C, Tu M, Wei L, Hsu T, Kao C, Chen T, Huang T. Calcium Silicate/Chitosan-Coated Electrospun Poly (lactic acid) Fibers for Bone Tissue Engineering. Preprints, 2017.

Subashri V, Nivedha V, Sherwood A, Abbott P, Gutmann J, Endo C, Farooq O, Aarthi M. Effect of laser assisted local anesthesia in single-visit root canal treatment for mandibular molar teeth with acute irreversible pulpitis. medRxiv, 2021.

Subramanya A, Prabhuji M. Dental practice in Pre-COVID19 and Future Perspectives. medRxiv, 2020.

Sudhakara P, Sellamuthu I, Aruni A. Bacterial Sialoglycosidases in Virulence and Pathogenesis. Preprints, 2019.

Sudhakara P, Gupta A, Bhardwaj A, Wilson A. Oral Dysbiotic Communities and Their Implications in Systemic Diseases. Preprints, 2018.

Sun H, Yu W, Sun S, Lee S, Farhood V, Afzali P. Characterization, Management, and Epidemiology of Odontogenic Infections: An Analysis of 103 Cases at a Major Regional Medical Center. Preprints, 2020.

Sunandhakumari V, Vidhyadharan A, Murali N, Alim A, J S, Shankar K. Root Membrane Technique-An Insight. Preprints, 2020.

Tada A, Miura H. Relationship between Vitamin C and Periodontal Diseases: A Systematic Review. Preprints, 2019.

Tanu L, Polii H, Lesmana D. Morphological Appearance of Condylar Head in Mixed Dentition Period (Evaluated from Panoramic Radiograph). Preprints, 2021.

Tefera A, Asefaw K, Bekele B, G/Mariam A, Ayelign A, Aragie H, Ayhualem S, Akalu Y, Molla M, Muche A. Dental Professionals Knowledge, Attitude, and Practice Towards to COVID-19: Systematic Review and Meta-Analysis. Preprints, 2020.

Terzi O, Yılmaz F. The Rational Drug Use of Dentists in a University Hospital. Preprints, 2018.

Thurzo A, Strunga M, Havlínová R, Reháková K, Urban R, Surovková J, Kurilová V. Smartphone-Based Facial Scanning as a Viable Tool for Facially Driven Orthodontics? Preprints, 2022.

Thurzo A, Bražinová A, Markovská N, Waczulíková I, Smatana M, Urbanová W, Strunga M, Kurilová V, Kaiferová J, Sabaka P. Biosafety in Dental Care Facing Highly Transmissible SARS-CoV-2 Variants: Introduction of Prospective Setting Protocol in Prevention of COVID-19. Preprints, 2022.

Thurzo A, Jančovičová V, Hain M, Thurzo M, Novák B, Kosnáčová H, Lehotská V, Varga I, Moravanský N. Human Remains Identification Using Micro-CT, Chemometric and A.I. Methods in Forensic Experimental Reconstruction of Dental Patterns after Concentrated Acid Significant Impact. Preprints, 2022.

Thurzo A, Urbanová W, Novák B, Waczulíková I, Varga I. 3D Printed Orthodontic Distalizer with Individual Base for Tooth-Borne Hybrid Approach in Class II Unilateral Malocclusions Treatment. Preprints, 2021.

Tomov G, Voynov P, Bachurska S. Granulomatous Cheilitis Revealing Latent Tuberculous Infection Preprints, 2022.

Tranby E, Frantsve-Hawley J, Minter-Jordan M, Thommes J, Jacob M, Monopoli M, Okunev I, Boynes S. A Cross-Sectional Analysis of Oral Health Spending Over the Lifespan in Commercial and Medicaid Insured Populations. medRxiv, 2021.

Tsukinoki K, Yamamoto T, Saito J, Sakaguchi W, Iguchi K, Inoue Y, Ishii S, Sato C, Yokoyama M, Shiraishi Y, Kato N, Shimada H, Makabe A, Saito A, Tanji M, Nagaoka I, Saruta J, Yamaguchi T, Kimoto S, Yamaguchi H. Prevalence of Salivary IgA Reacting with SARS-CoV-2 among Japanese People Unexposed to the Virus. medRxiv, 2022.

Tsukinoki K, Yamamoto T, Handa K, Iwamiya M, Saruta J, Ino S, Sakurai T. Detection of cross-reactive IgA against SARS-CoV-2 spike 1 subunit in saliva. medRxiv, 2021.

Vasudevan K, Stahl V. CBD-Supplemented Polishing Powder Enhances Tooth Polishing by Inhibiting Dental Plaque Bacteria. Preprints, 2020.

Vernon J, Black E, Dennis T, Devine D, Fletcher L, Wood D, Nattress B. Dental mitigation strategies to reduce aerosolization of SARS-CoV-2. medRxiv, 2021.

Vintanel-Moreno C, Martínez-González J, Martínez-Rodríguez N, Meniz-García C, Leco-Berrocal I. Implant Surgery. Analysis of Adverse Effects and Haemodynamic Changes. Preprints, 2021.

Wang D, Tsai F, Tu H, Yang C, Hsu M, Huang L, Lin C, Hsu W, Lin Y. Effects of nasogastric tube on oral microbiome among long-term care patients. medRxiv 2022.

Wang F, Liu D, Zhuang Y, Feng B, Lu W, Yang J, Zhuang G. Mendelian randomization analysis identified genes potentially pleiotropically associated with periodontitis. medRxiv, 2021.

Wilde D, Kansara S, Sapienza L, Banner L, Morlen R, Hernandez D, Huang A, Mai W, Fuller C, Lai S, Sandulache V. Early detection of mandible osteoradionecrosis risk in a high comorbidity Veteran population. medRxiv, 2022.

Xu F, Aboseria E, Janal M, Pushalkar S, Bederoff M, Vasconcelos R, Sapru S, Paul B, Queiroz E, Makwana S, Solarewicz J, Guo Y, Aguallo D, Gomez C, Shelly D, Aphinyanaphongs Y, Gordon T, Corby P, Kamer A, Li X, Saxena D. Comparative Effects of E-cigarette Aerosol on Periodontium. medRxiv, 2021.

Yoshino Y, Miyaji H, Nishida E, Kanemoto Y, Hamamoto A, Kato A, Sugaya T, Akasaka T. Periodontal Tissue Regeneration by Recombinant Human Collagen Peptide Granules Applied With β-Tricalcium Phosphate Fine Particles. Preprints, 2022.

Zhang Y, Rocca J, Fornaini C, Zhen Y, Zhao Z, Merigo E. Er:YAG Laser Debonding of Porcelain Laminate Veneers. Preprints, 2018.

Zhang H, Shan J, Zhang P, Chen X, Jiang H. Trabeculae microstructure parameters serve as effective predictors for marginal bone loss of dental implant in the mandible. medRxiv, 2020.

Ziebart T, Blatt S, Jung J, Pabst A, Walter C, Halling F, Heymann P, Neff A, Righesso L. Impact of Soft Tissue Pathophysiology in the Development and Maintenance of Bisphosphonate Related Osteonecrosis of the Jaw (BRONJ). Preprints, 2016.

